# Implementation of an in-house real-time reverse transcription-PCR assay for the rapid detection of the SARS-CoV-2 Marseille-4 variant

**DOI:** 10.1101/2021.02.03.21250823

**Authors:** Marielle Bedotto, Pierre-Edouard Fournier, Linda Houhamdi, Anthony Levasseur, Jeremy Delerce, Lucile Pinault, Abdou Padane, Amanda Chamieh, Hervé Tissot-Dupont, Philippe Brouqui, Cheikh Sokhna, Eid Azar, Rachid Saile, Souleymane Mboup, Idir Bitam, Philippe Colson, Didier Raoult

**Affiliations:** IHU Méditerranée Infection, 19-21 boulevard Jean Moulin, 13005 Marseille, France; Aix-Marseille Univ., Institut de Recherche pour le Développement (IRD), Assistance Publique - Hôpitaux de Marseille (AP-HM), Microbes Evolution Phylogeny and Infections (MEPHI), 27 boulevard Jean Moulin, 13005 Marseille, France; Institut de Recherche en Santé, de Surveillance Épidémiologique et de Formations (IRESSEF), arrondissement 4 rue 2D1, pôle urbain de Diamniadio, Dakar, Sénegal; Saint George Hospital University Medical Center, University of Balamand, Beirut, Lebanon; Vecteurs - Infections Tropicales et Méditerranéennes (VITROME), Campus International IRD-UCAD de l’IRD, Dakar, Senegal; Aix-Marseille Univ., Institut de Recherche pour le Développement (IRD), Assistance Publique - Hôpitaux de Marseille (AP-HM), Vecteurs - Infections Tropicales et Méditerranéennes (VITROME), 27 boulevard Jean Moulin, 13005 Marseille, France; Hassan II University of Casablanca, Morocco; Ecole supérieure en sciences de l’aliment et des industries agro-alimentaires, Alger, Algeria

**Author notes:** **Corresponding author:** Didier RAOULT, IHU - Méditerranée Infection, 19-21 boulevard Jean Moulin, 13005 Marseille, France.

**Keywords:** SARS-CoV-2, Covid-19, variant, Marseille-4, qPCR, diagnosis, molecular epidemiology

## Abstract

**Introduction:** The SARS-CoV-2 pandemic has been associated with the occurrence since summer 2020 of several viral variants that overlapped or succeeded each other in time. Those of current concern harbor mutations within the spike receptor binding domain (RBD) that may be associated with viral escape to immune responses. In our geographical area a viral variant we named Marseille-4 harbors a S477N substitution in this RBD.

**Materials and methods:** We aimed to implement an in-house one-step real-time reverse transcription-PCR (qPCR) assay with a hydrolysis probe that specifically detects the SARS-CoV-2 Marseille-4 variant.

**Results:** All 6 cDNA samples from Marseille-4 variant strains identified in our institute by genome next-generation sequencing (NGS) tested positive using our Marseille-4 specific qPCR, whereas all 32 cDNA samples from other variants tested negative. In addition, 39/42 (93%) respiratory samples identified by NGS as containing a Marseille-4 variant strain and 0/26 samples identified as containing non-Marseille-4 variant strains were positive. Finally, 1,585/2,889 patients SARS-CoV-2-diagnosed in our institute, 10/277 (3.6%) respiratory samples collected in Algeria, and none of 207 respiratory samples collected in Senegal, Morocco, or Lebanon tested positive using our Marseille-4 specific qPCR.

**Discussion:** Our in-house qPCR system was found reliable to detect specifically the Marseille-4 variant and allowed estimating it is involved in more than half of our SARS-CoV-2 diagnoses since December 2020. Such approach allows the real-time surveillance of SARS-CoV-2 variants, which is warranted to monitor and assess their epidemiological and clinical characterics based on comprehensive sets of data.

## 1. Introduction

Since the onset of the SARS-CoV-2 pandemic in December 2019 in China, almost 100 million cases have been reported worldwide as on January 28^th^, 2021 (https://swww.ecdc.europa.eu/en/covid-19-pandemic). This has been associated with the occurrence since summer 2020 of several viral variants that overlapped or succeeded each other in time [1-3]. Those of current concern harbor mutations within the spike glycoprotein, particularly within the spike receptor binding domain (RBD) that leads to viral entry into host cells by binding to the ACE2 receptor (Figure 1) [4]. Such SARS-CoV-2 variants include the 20I/501Y.V1 [3], 20H/501Y.V2 [5], and 20J/501Y.V3 [6] strains that harbor a N501Y substitution in the spike RBD and were reported in the UK and in South Africa, as highly transmissible, and in Brazil, respectively. In our geographical area we detected 10 viral variants since June 2020 [1]. One of them, we named Marseille-4, harbors a S477N substitution in the spike RBD that has been associated with an improved binding affinity to ACE2 [6] and a broad resistance to monoclonal neutralizing antibodies [7]. It predominates in Marseille since August 2020, has been reported to spread in Europe since early summer and was classified as the Nextstrain 20A.EU2 lineage [1, 2]. The continuous emergence of new SARS-CoV-2 variants, including some of substantial concern regarding their transmissibility and their possible ability to evade immune responses [8-10], warrants to set up strategies for their detection and surveillance. SARS-CoV-2 incidence is currently substantial in several countries including France, and in our institute we for instance diagnose >100 new cases daily. Therefore, alternative strategies to sequencing are useful for variant screening. We aimed to implement an in-house one-step real-time reverse transcription-PCR (qPCR) assay that specifically detects the SARS-CoV-2 Marseille-4 variant.

**Fig. 1.**
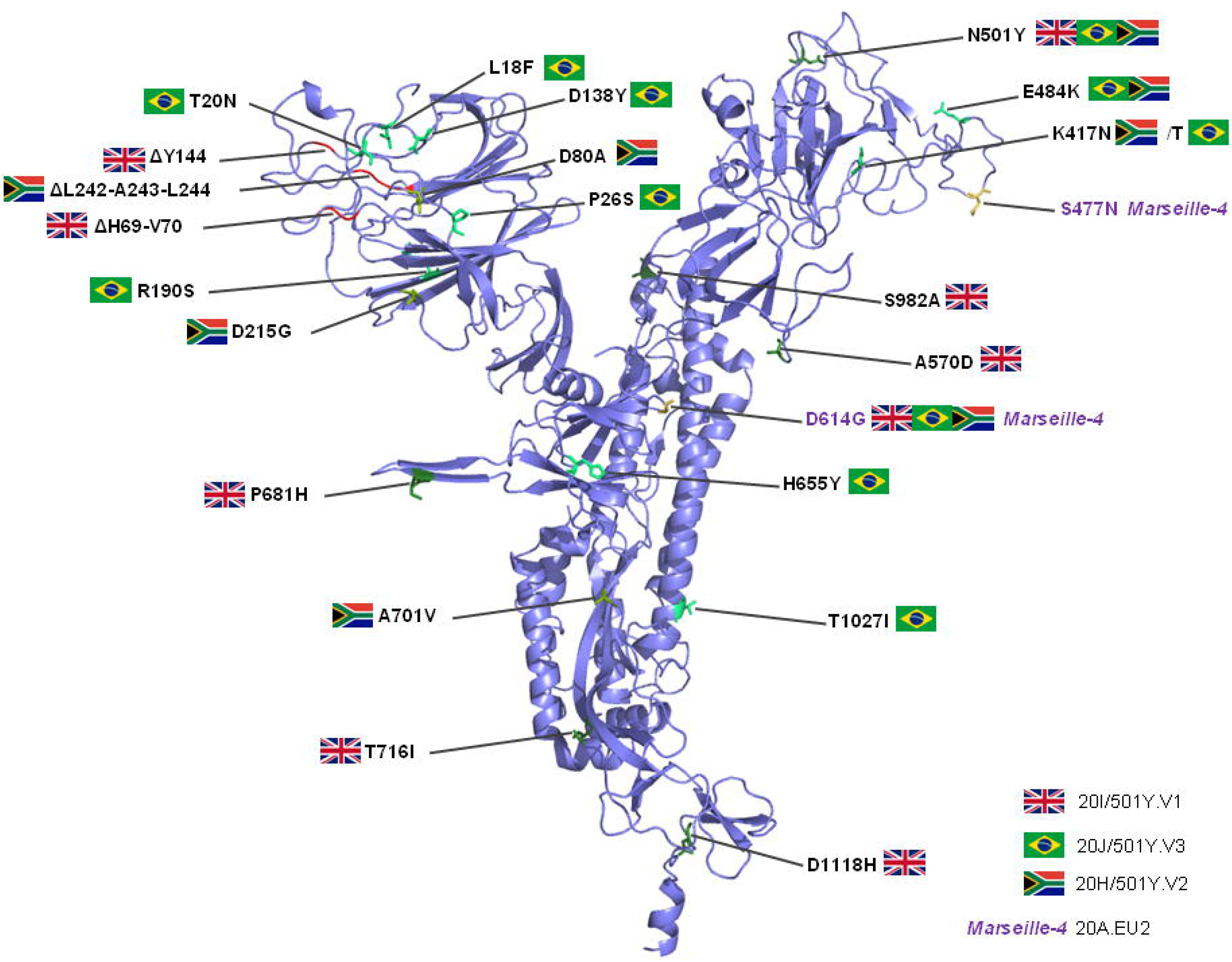

## 2. Material and methods

SARS-CoV-2 genomes from our institute sequence database and from the GISAID database (https://www.gisaid.org/) were used to design a primer pair and a hydrolysis probe. These sequences target a fragment of the nsp4 gene that contains nucleotide position 9,526 of the viral genome [in reference to genome GenBank Accession no. NC_045512.2 (Wuhan-Hu-1 isolate)] where is located a hallmark mutation G>U of the SARS-CoV-2 Marseille-4 variant. The sequences of the qPCR primers and probe are shown in Table 1. The qPCR was performed by adding 5 μL of extracted viral RNA to 15 μL of reaction mixture containing 5 μL of 4X TaqMan Fast Virus 1-Step Master Mix (Thermo Fisher Scientific, Grand Island, NY, USA), 0.5 μL of forward primer (10 pmol/µL), 0.5 μL of reverse primer (10 pmol/µL), 0.4 μL of probe (10 pmol/µL), and 8.6 μL of water. PCR conditions were as follows: reverse transcription at 50°C for 10 min, then a hold at 95°C for 20 sec followed by 40 cycles comprising a denaturation step at 95°C for 15 sec and a hybridization-elongation step at 60°C for 60 sec. This qPCR was run on a LC480 thermocycler (Roche Diagnostics, Mannheim, Germany).

**Table 1.**
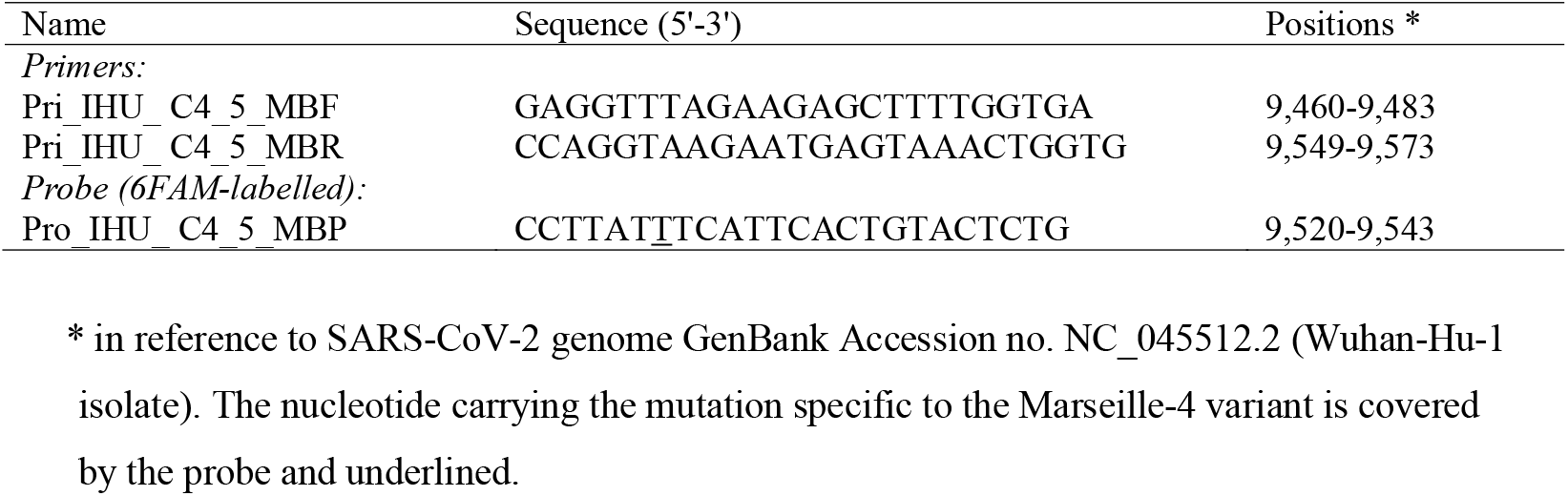
Primers and probe of the Marseille-4 variant-specific Qpcr

## 3. Results

Firstly, we tested a panel of 38 cDNA samples from each of the 10 variants named Marseille-1 to Marseille-10 that we identified by genome next-generation sequencing (NGS) and circulated since summer 2020 in our geographical area (6 from Marseille-4 strains, 5 from Marseille-5 strains, 5 from Marseille-3 strains, 4 from Marseille-1 strains, and 3 from strains classified in each of the variants Marseille-2, Marseille-6, Marseille-7, Marseille-8, Marseille-9, and Marseille-10) [1]. All 6 Marseille-4 samples tested positive whereas all 32 samples from other variants tested negative. Secondly, we tested 42 samples identified in our institute by genome NGS [1] as being from patients infected with a SARS-CoV-2 Marseille-4 variant: 39 of them (93%) were positive using our Marseille-4 specific qPCR. Thirdly, we tested 26 samples identified by next-generation genome sequencing as containing SARS-CoV-2 strains that were not Marseille-4 variants (including 17 N501Y variants, 5 Marseille-2 variants, 3 clade 20A strains and 1 clade 20C strain): none of them were positive using our Marseille-4 specific qPCR. Positive and negative predictive values of Marseille-4 detection by our qPCR were 100% and 90%, respectively. Finally, we tested with our Marseille-4 specific qPCR the respiratory samples from 2,889 patients SARS-CoV-2-diagnosed in our institute. None of 22 patients’ samples collected in June, 20 (5.6%) of 357 patients’ samples collected in July, and 1,565 (53%) of 2,954 patients’ samples collected in December and January tested positive (Figure 2). These results are congruent with those obtained based on genome NGS that showed that the Marseille-4 variant emerged in our geographical area in July and has been predominant since August 2020 [1]. In addition, we found that 10 (3.6%) of 277 respiratory samples collected in Algeria in September-October tested positive using our Marseille-4 specific qPCR, while none of 94 respiratory samples collected in Senegal in September-October, of 94 samples collected in Morocco in November 2020, and of 19 samples collected in Lebanon in October 2020, tested positive.

**Fig. 2.**
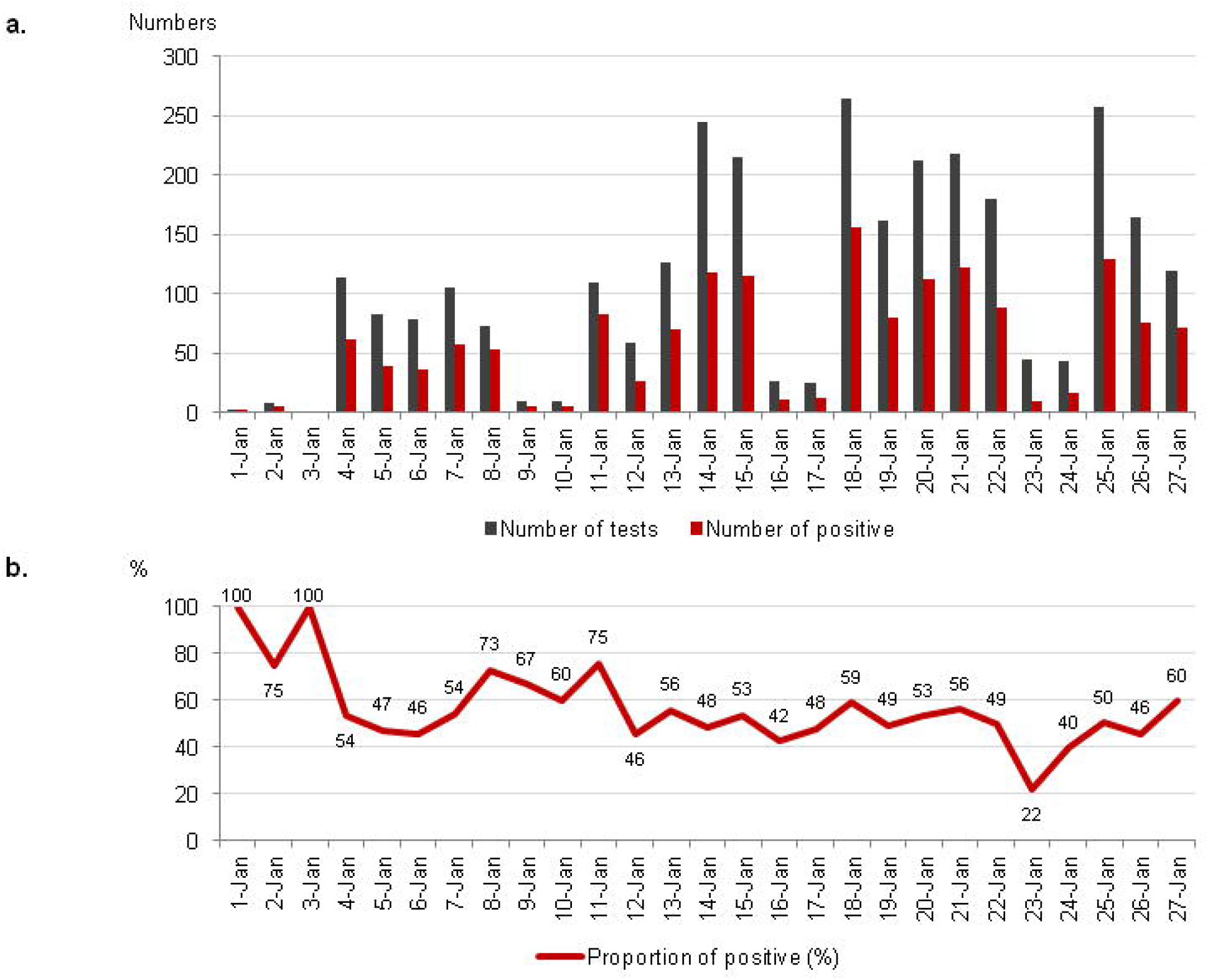

## 4. Discussion

Our in-house qPCR system was found reliable to detect specifically the Marseille-4 variant and allowed estimating it is involved in more than half of our SARS-CoV-2 diagnoses since December 2020. This assay is currently routinely used in our clinical microbiology and virology laboratory to screen systematically all samples found SARS-CoV-2-positive using the first-line qPCR diagnosis assay, which allows the real-time classification of viral strains in more than half of the diagnoses (Figure 2). In case of negativity of this Marseille-4 specific qPCR, samples are tested using alternative qPCR assays that are specific to other variants that circulate at a lower incidence level than the Marseille-4 variant, or they are submitted to next-generation sequencing in case of cycle threshold value (Ct) <18 with the SARS-CoV-2 qPCR diagnosis test [1, 11]. Such approach based on qPCR assays targeting specifically SARS-CoV-2 variants allows their real-time surveillance, which is warranted to monitor and assess their epidemiological and clinical characterics based on comprehensive sets of data. In addition, in-house qPCR assays can be implemented rapidly, easily and at low cost on various open qPCR microplate platforms, which may allow adapting continuously the diagnosis strategies to the emergence and dynamics of SARS-CoV-2 variants.

## Data Availability

Data underlying the study are available from the corresponding author upon request.

## Credit authorship contribution statement

Conceived and designed the experiments: DR, PC. Contributed materials/analysis tools: MB, PEF, LH, AL, JD, LP, AP, AC, HTD, PB, CS, EA, RS, SM, IB, PC. Analyzed the data: MB, PC, PEF, DR. Wrote the paper: PC, MB, DR.

## Ethics

This study has been approved by the ethics committee of the University Hospital Institute Méditerranée Infection, Marseille, France, with the registration number 2020-029.

## Acknowledgments

This manuscript text has been edited by a native English speaker.

